# Prevalence and associated factors of non-communicable diseases among men in Kenya: an analysis of the 2022 Kenya Demographic and Health Survey

**DOI:** 10.1101/2025.06.13.25329608

**Authors:** John Baptist Asiimwe, Lilian Nuwabaine, Angella Namulema, Quraish Sserwanja, Joseph Kawuki, Grace Nambozi

## Abstract

**Background:** Non-communicable diseases (NCDs) are the leading cause of morbidity and mortality worldwide, with low- and middle-income countries (LMICs) increasingly being disproportionately affected. Although anecdotal evidence or reports indicate an increasing number of Kenyan men having NCDs, the prevalence and associated factors are not well understood. Therefore, this study aimed to determine the prevalence and associated factors of non-communicable diseases among men in Kenya.

**Methods:** Secondary data comprising 14,453 men aged 15-54 years from the 2022 Kenya Demographic and Health Survey (KDHS) were analysed using univariable and multivariable logistic regression analyses in SPSS, version 29.

**Results:** Overall, the percentage of men with at least one NCD was 9.4% (95% confidence interval [CI]: 8.7-10.2%). Whereas the proportion of participants with multiple NCDs was 1.9 (95CI:1.6-2.3). Across NCDs, the highest prevalent NCD was hypertension (3.5% 95%CI:3.1-3.9) followed by depression (2.2% (95%CI:1.9-2.5), anxiety (1.6% (95%CI:1.3-1.9), arthritis (1.4% (95%CI:1.1-1.6), heart disease (1.2% (95%CI:0.9-1.5), diabetes (1% (95%CI:0.8-1.3), lung disease (1% (95%CI:0.7-1.3), and cancer (0.1% (95%CI:0-0.1). In terms of multiple chronic conditions (multimorbidity), the majority of the participants had diabetes and hypertension (0.5% (95%CI:0.3-0.7) followed by hypertension and depression (0.3% (95%CI:0.2-0.4), hypertension and anxiety (0.3% (95%CI:0.2-0.4), and arthritis and depression (0.2% (95%CI:0.1-0.2). Several factors, such as age, region, residence, ethnicity, education level, health status, wealth index, religion, media access, living a sedentary lifestyle, and physical activity, were found to be significantly associated with the prevalence of NCDs.

**Conclusion:** The overall prevalence of NCDs among men is relatively lower than that of Kenyan women. We found that sociodemographic and lifestyle factors were significantly associated with the prevalence of NCDs. To reduce non-communicable diseases (NCDs) among men, tailored health education, medical checkups, and physical activity promotion are essential. Region-specific policies and culturally sensitive interventions should address risk factors and ethnic disparities. Wealthier, more educated men should be encouraged to adopt healthier lifestyles, while rural areas need improved access to specialized healthcare services. The media should balance health messaging, and religious communities/leaders can play a supportive role in promoting health and reducing stigma.

## Introduction

Non-communicable diseases (NCDs) are a significant global health challenge, contributing to the top ten causes of death across all income levels [1]. The World Health Organization (WHO) reports that cardiovascular diseases are the leading cause of NCD-related deaths, followed by cancers, chronic respiratory diseases, and diabetes. Low Middle middle-income countries (LMICs) account for 77% of these deaths [2]. In 2021, NCDs were responsible for at least 43 million deaths, equivalent to 75% of non-pandemic-related deaths globally, and 73% of the deaths were in low- and middle-income countries [3].

In Sub-Saharan Africa (SSA), nearly 30% of NCD deaths occur in individuals under 60 years, compared with 13% in high-income countries [4]. The prevalence of NCDs in SSA has risen steadily, with disability-adjusted life-years (DALYs) associated with NCDs increasing by 67% from 1990 to 2017 [5, 6]. Kenya, with a population of 49% male and 51% female, has a life expectancy of 64.7 years for men and 69 years for women (KNBS, 2022). Kenya is transitioning from a focus on communicable diseases, such as malaria, HIV/AIDS, and tuberculosis, to a growing burden of NCDs due to behavioral changes and rural-urban migration [7]. Common NCDs in Kenya include stroke, coronary heart disease, dementia, and certain cancers [8].

The rise of NCDs can be attributed to multiple factors, with behavioral changes being a primary driver [9]. Risk factors such as smoking, physical inactivity, obesity, and hypertension are prevalent in both high-income and low- and middle-income countries, including Kenya and SSA[10]. These risk factors are exacerbated by urbanization, with increased exposure to unhealthy diets, sedentary lifestyles, and alcohol consumption contributing significantly to the rise of NCDs [6]. In Kenya, the urban poor population is particularly vulnerable, as they face increased exposure to these common risk factors [4, 11]. Furthermore, gender plays a role in the prevalence of NCDs, as men tend to engage in higher-risk behaviors such as smoking and excessive alcohol consumption, which increase their vulnerability to diseases like cardiovascular conditions and diabetes [12].

Kenya has implemented several measures to address the growing NCD burden. The Non-Communicable Diseases and Injury (NCDI) Poverty Commission in Kenya is tasked with assessing disease burden and identifying health service gaps while prioritizing cost-effective interventions [8]. The country’s response also includes the Kenya National Strategy for the Prevention and Control of Non-Communicable Diseases (2015–2020), which aims to reduce NCD-related mortality through prevention, early detection, and healthcare improvements. Despite these efforts, challenges persist, particularly in the allocation of sufficient resources and funding to NCD interventions. Limited prioritization and financial support hinder the full implementation of strategies designed to address NCDs. In 2018, 37% of DALYs lost in Kenya were attributed to NCDs, underscoring their serious public health impact [13]. The inclusion of NCDs in the Kenya Demographic and Health Survey (KDHS) signals recognition of their growing importance, but there remain gaps in comprehensive data collection and monitoring, which hamper effective policy-making. Therefore, this study aimed to determine the prevalence and associated factors of non-communicable diseases among men in Kenya.

## Methods

### Data source, sample design, and collection

This study made use of the 2022 Kenya Demographic and Health Survey (KDHS), which used a two-stage stratified sampling approach. The initial stage included the selection of 1,692 enumeration areas (EAs) or clusters from a master sampling frame comprising 129,067 EAs based on the 2019 Kenya population and housing census, employing equal probability and independent selection[14]. The house listing was then created to establish a sampling frame, which was utilized in the next stage to choose 25 households from each cluster. However, if a cluster had fewer than 25 households, all of them were included in the sample. Ultimately, the survey was carried out in 1691 clusters. The Inner-City Fund (ICF) assisted in the pretesting of the study instruments and in training the data collectors, with data collection occurring between February and July of 2022. All men aged 15-54 years who were regular members of the chosen households or who had spent the previous night in those homes were interviewed in either Swahili or English. [14]. Out of 14,818 men who responded to the survey, 14, 453 were included in this analysis (98% response rate) [14]. The 2022 KDHS dataset was requested, and we secured written permission to use it from the MEASURE DHS website (https://www.dhsprogram.com/data/available-datasets.cfm). While the dataset includes numerous variables, we focused solely on those that were pertinent and applicable to our research.

### Study variables

#### Dependent/outcome

The main outcome of this study was NCDs. The study analysed the prevalence and factors associated with eight NCDs that were self-reported by the DHS participants and included heart disease, diabetes, hypertension, arthritis, anxiety, lung disease, depression, and cancer. During the survey, participants were asked to mention whether a doctor or other healthcare workers had informed them of having any of above mentioned NCDs. Men who answered yes to any NCD were coded as 1; otherwise, as 0. Two composite variables were constructed as secondary outcomes of the study, and these included having at least 1 NCD (1NCD/yes vs none/no) and having multiple NCDs (≥2NCDs (yes) vs ≤1 (No).

#### Independent variables

The factors included in the analysis were categorized into two namely sociodemographic and lifestyle behavioural factors. The 12 sociodemographic factors that were included in the analysis, namely; ethnicity (categorized into Kenya’s 12 major tribes; Maasai, Mijikenda/Swahili, Somali, Meru, Taita/Taveta, Embu, Kalenjin, Kamba, Kikuyu, Kisii, Luhya, Luo, and Others), region (categorized into Kenya’s eight provinces; Northeastern, Central, Western, Nairobi, Nyanza, Rift Valley, Eastern, and Coast), religion (Christian, Muslim, or others), marital status (unmarried or married/cohabiting), working status (Yes vs No), household size (≤4 vs ≥5 members), education (tertiary, secondary or none/primary), residence (urban vs rural), age in years (35-54, 25-34, 15-24), and wealth index (richest, richer, middle, poorer, and poorest). The wealth index was calculated by the 2022 KDHS from information on household asset ownership using principal component analysis [14]. The participants’ perceived health status at the time of the interview (good, moderate, bad), and exposure to mass media outlets like newspapers, the internet, television, and radio (Yes vs No) were also considered in the analysis.

Five lifestyle behavioural factors were also analysed: whether the participants consumed alcohol, used tobacco (yes/no), including smoked or smokeless tobacco, or had a history of smoking tobacco. The number minutes one exercised per week (categorised as adequate (≥150 minutes vs inadequate (<150 minutes) and the number hours one spent seated (grouped as high risk (>8 hours)/sedentary life vs Low risk (≤8 hours)/non sedentary) were also included in the analysis.

### Statistical analysis

The data was analyzed using the complex samples package in SPSS (V29), which addressed the complex sample design inherent in DHS data.[15, 16]. The complex sample package delivers accurate estimates of parameters since it considers sample weighting, clustering, and stratification that took place during the selection of study participants.[15, 16]. Furthermore, to address the unequal sampling probabilities across various strata and to guarantee that the study results are representative, sample weights from the DHS were utilized for all computed frequencies [15, 16]. Prior to the analysis, the data underwent cleaning, and dummy variables were generated. Descriptive statistics, including frequencies, were calculated for all categorical variables at the univariate level. Univariate and multivariate logistic regression analyses were conducted to identify independent factors related to the prevalence of NCDs. All variables with P-values less than 0.05 were included into a simple multivariate logistic regression to determine the factors associated with the prevalence of NCDs. All odds ratios are presented with their 95% confidence intervals. Additionally, multi-collinearity among all predictor variables in the model was evaluated using a variance inflation factor (VIF), with a threshold of greater than 10 considered significant. [15, 16]. All of the factors fell below the threshold.

### Ethical consideration

No ethical clearance was necessary to examine the secondary data since it is accessible to the public. Nevertheless, authorization to use the 2022 KDHS datasets was secured from MEASURE DHS (https://www.dhsprogram.com/data/available-datasets.cfm). Ethical permission for the research detailed in the datasets was granted by the ICF Institutional Review Board. The Kenya National Bureau of Statistics carried out the study in collaboration with several other partners. Written informed consent was obtained from human participants and written informed consent was also obtained from legally authorized representatives of minor participants.

## Results

### Socio-demographic Characteristics of the study participants

In total, 14,453 men were included in this analysis (Table 1). Most of the participants were aged 15-34 years (66.7%), lived in the rural setting (61.1%), identified as being from the Rift Valley, Eastern, Central, and Nairobi provinces (67.3%). The majority identified as being from the Kikuyu, Luhya, and Kalenjin, and Kamba tribes of Kenya (58.3%), were Christians by faith (85.5%), unmarried (51.9%), working (78.2%), and had completed utmost secondary education (79.7%). In total, 65.8% of the participants belonged to the middle, richer, and richest quintiles, and 95% lived in households with more than five members. The majority were exposed to mass media, which included radio (87.4%), television (80.1%), the internet (58.8%), and newspapers (39.4%), and described themselves as being in good health conditions (84.1%). In terms of lifestyle behaviours, 26.6% of the participating men consumed alcohol, 13.3% used tobacco, and 41.7% had smoked tobacco in the past. Additionally, the majority (80.5%) of the participants had adequate weekly exercise and lived a non-sedentary lifestyle (94.8%).

**Table 1.** Socio-demographic Characteristics of the study participants

| Variable | *n* | Weighted % |
| --- | --- | --- |
| **Age (years)** |  |  |
| 15-24 | 5,579 | 38.6 |
| 25-34 | 4,055 | 28.1 |
| 35-59 | 4,819 | 33.3 |
| **Residence** |  |  |
| Urban | 5,624 | 38.9 |
| Rural | 8,829 | 61.1 |
| **Region/Province** |  |  |
| Coast | 1,341 | 9.3 |
| Northeastern | 273 | 1.9 |
| Eastern | 2,123 | 14.7 |
| Central | 1,945 | 13.5 |
| Rift Valley | 3,808 | 26.3 |
| Western | 1,499 | 10.4 |
| Nyanza | 1,610 | 11.1 |
| Nairobi | 1,855 | 12.8 |
| **Ethnicity** |  |  |
| Embu | 186 | 1.3 |
| Kalenjin | 1,823 | 12.6 |
| Kamba | 1,669 | 11.5 |
| Kikuyu | 2,521 | 17.4 |
| Kisii | 901 | 6.2 |
| Luhya | 2,424 | 16.8 |
| Luo | 1,591 | 11.0 |
| Maasai | 325 | 2.2 |
| Meru | 717 | 5.0 |
| Mijikenda/Swahili | 798 | 5.5 |
| Somali | 294 | 2.0 |
| Taita/Taveta | 136 | 0.9 |
| Other | 1,068 | 7.4 |
| **Education** |  |  |
| None/primary | 5,660 | 39.2 |
| Secondary | 5,851 | 40.5 |
| Tertiary | 2,943 | 20.4 |
| **Religion** |  |  |
| Christians | 12,361 | 85.5 |
| Muslims | 1,042 | 7.2 |
| Others | 1,050 | 7.3 |
| **Marital status** |  |  |
| Unmarried (single divorced/widowed/separated) | 7,495 | 51.9 |
| Married/Cohabiting | 6,958 | 48.1 |
| **Working status/occupation** |  |  |
| Not Working | 3,157 | 21.8 |
| Working | 11,296 | 78.2 |
| Wealth index |  |  |
| Poorest | 2,175 | 15.0 |
| Poorer | 2,762 | 19.1 |
| Middle | 2,932 | 20.3 |
| Richer | 3,504 | 24.2 |
| Richest | 3,079 | 21.3 |
| **Household number** |  |  |
| ≤4 | 717 | 5.0 |
| ≥5 | 13,736 | 95.0 |
| **Health status** |  |  |
| Bad | 189 | 1.3 |
| Moderate | 2,116 | 14.6 |
| Good | 12,148 | 84.1 |
| **Newspapers** |  |  |
| No | 8,753 | 60.6 |
| Yes | 5,700 | 39.4 |
| **TV** |  |  |
| No | 2,875 | 19.9 |
| Yes | 11,578 | 80.1 |
| **Radio** |  |  |
| No | 1,820 | 12.6 |
| Yes | 12,633 | 87.4 |
| **Internet use** |  |  |
| No | 5,957 | 41.2 |
| Yes | 8,496 | 58.8 |
| **Tobacco use** |  |  |
| No | 12,525 | 86.7 |
| Yes | 1,928 | 13.3 |
| **Past smoking** |  |  |
| No | 288 | 58.3 |
| Yes | 206 | 41.7 |
| **Lifestyle Behavioural factors** |  |  |
| **Exercise Minutes per week** |  |  |
| Adequate (≥150 minutes) | 10,043 | 80.5 |
| Inadequate (<150 minutes) | 2,426 | 19.5 |
| **Number of hours per day seated/sedentary** |  |  |
| Low risk (≤8 hours)/non sedentary | 13,692 | 94.8 |
| High risk (>8 hours)/sedentary life | 747 | 5.2 |
| **Alcohol consumption** |  |  |
| No | 10,608 | 73.4 |
| Yes | 3,845 | 26.6 |

### Prevalence of NCDs among men in Kenya

Overall, the percentage of men with at least one NCD was 9.4% (95%CI:8.7-10.2%, Table 2). Whereas the proportion of participants with multiple NCDs was 1.9 (95CI:1.6-2.3). Across NCDs, the highest prevalent NCD was hypertension (3.5% 95%CI:3.1-3.9) followed by depression (2.2% (95%CI:1.9-2.5), anxiety (1.6% (95%CI:1.3-1.9), arthritis(1.4% (95%CI:1.1-1.6), heart disease(1.2% (95%CI:0.9-1.5), diabetes(1% (95%CI:0.8-1.3), lung disease(1% (95%CI:0.7-1.3), and cancer (0.1% (95%CI:0-0.1). In terms of multimorbidity, the majority of the participants had diabetes and hypertension (0.5 (95%CI:0.3-0.7) followed by hypertension and depression (0.3 (95%CI:0.2-0.4), hypertension and anxiety (0.3 (95%CI:0.2-0.4) and arthritis and depression (0.2 (95%CI:0.1-0.2).

**Table 2.** Prevalence of NCDs among men in Kenya

| Variable (N= 14453) | *n* | Weighted (95CI) |
| --- | --- | --- |
| Hypertension | 504 | 3.5 (3.1-3.9) |
| Diabetes | 144 | 1 (0.8-1.3) |
| Heart disease | 172 | 1.2 (0.9-1.5) |
| Lung disease | 139 | 1 (0.7-1.3) |
| Depression | 319 | 2.2 (1.9-2.5) |
| Anxiety | 227 | 1.6 (1.3-1.9) |
| Arthritis | 197 | 1.4 (1.1-1.6) |
| Cancer | 8 | 0.1 (0-0.1) |
| Living with atleast 1NCD | 1361 | 9.4 (8.7-10.2) |
| Living with $\geq 2$ NCDs | 14174 | 1.9 (1.6-2.3) |
| **Multi-morbidity** |  |  |
| Hypertension & Diabetes | 68 | 0.5 (0.3-0.7) |
| Hypertension & Depression | 47 | 0.3 (0.2-0.4) |
| Diabetes & Depression | 11 | 0.1 (0.041-0.1) |
| Hypertension & Anxiety | 39 | 0.3 (0.2-0.4) |
| Diabetes & Anxiety | 12 | 0.1 (0.04-0.2) |
| Arthritis & Depression | 23 | 0.2 (0.1-0.2) |
| Arthritis And Anxiety | 8 | 0.1 (0.03-0.1) |
| Heart Disease & Depression | 12 | 0.1 (0.05-0.2) |
| Heart Disease & Anxiety | 6 | 0.04 (0.02-0.1) |
| Diabetes & Hypertension & Depression | 8 | 0.1 (0.02-0.1) |
| Diabetes & Hypertension & Anxiety | 9 | 0.1 (0.02-0.2) |
| Diabetes & Hypertension & Depression & Anxiety | 3 | 0.02 (0.01-0.1) |

Note. NCD=Non communicable diseases. Due to a small sample size (n=8) of those who self-reported to have cancer further analysis of data related to cancer was not undertaken

### Factors associated with having specific/ multiple NCDs

Table 3 and 4 summarizes the results of univariable, and multivariable analysis of the factors associated with each specific NCD and having multiple NCDs, respectively. Several factors were found to be associated with having specific and multiple NCDs which included age, region, residence, ethnicity, education level, health status, wealth index, religion, media access, living sedentary lifestyle and inadequate exercises. We found that compared with younger men (15-24 years), older participants (≥25years) had a higher likelihood of experiencing hypertension (aOR 7.00 (95%CI: 3.75-13.08), diabetes mellitus (aOR 6.54 (95%CI: 2.72-16.22), arthritis (aOR 2.62 (95%CI:1.33-5.13), depression (aOR 5.61 (95%CI: 3.15-9.97), having atleast one NCD (aOR2.94 (95%CI:2.17-3.97) and multiple NCDs (aOR 6.95 (95%CI:2.66-18.19).

**Table 3.** Factors associated with having specific/ individual NCDs among men in Kenya

| Variable | n | Hypertension |  |  | Diabetes Mellitus |  |  | Heart diseases |  |  | Lung disease |  |  |
| --- | --- | --- | --- | --- | --- | --- | --- | --- | --- | --- | --- | --- | --- |
|  |  | Yes, % | uORs (95CI) | aORs (95CI) | Yes, % | uORs (95CI) | aORs (95CI) | Yes, % | uORs (95CI) | aORs (95CI) | Yes, % | uORs (95CI) | aORs (95CI) |
| **Age (years)** |  |  |  |  |  |  |  |  |  |  |  |  |  |
| 15-24 | 5579 | 0.32 | 1 | 1 | 0.06 | 1 | 1 | 0.57 | 1 | - | 0.40 | 1 | - |
| 25-34 | 4055 | 0.62 | **2.68**<br>(1.58-4.56)* | 1.78<br>(0.92-3.43) | 0.13 | **3.02**<br>(1.19-7.63)** | 1.23<br>(0.43-3.52) | 0.27 | 0.64<br>(0.34-1.22) | - | 0.18 | 0.62<br>(0.24-1.57) | - |
| 35-59 | 4819 | 2.53 | **9.62**<br>(6.30-14.70)* | **7.00**<br>(3.75-13.08)* | 0.81 | **16.12**<br>(7.91-32.88)** | **6.64**<br>(2.72-16.22)** | 0.35 | 0.71<br>(0.40-1.25) | - | 0.38 | 1.09<br>(0.51-2.35) | - |
| **Residence** |  |  |  |  |  |  |  |  |  |  |  |  | - |
| Urban | 5624 | 1.47 | 1.15<br>(0.87-1.51) | - | 0.45 | 1.31<br>(0.78-2.20) | - | 0.52 | 1.20<br>(0.69-2.10) | - | 0.49 | 1.63<br>(0.85-3.11) | - |
| Rural | 8829 | 2.02 | 1 | - | 0.54 | 1 | - | 0.68 | 1 | - | 0.47 | 1 | - |
| **Region/Province** |  |  |  |  |  |  |  |  |  |  |  |  |  |
| Coast | 1341 | 0.36 | 1 | - | 0.06 | 1 | 1 | 0.06 | 1 | 1 | 0.06 | 1 | 1 |
| Northeastern | 273 | 0.04 | 0.52<br>(0.26-1.03) | - | 0.02 | 1.92<br>(0.83-4.47) | 4.61<br>(0.96-22.08) | 0.01 | 0.53<br>(0.15-1.83) | **6.51**<br>(1.11-38.21)* |  | 0.00<br>(0.00-0.00) | 628(0.00-725) |
| Eastern | 2123 | 0.45 | 0.77<br>(0.52-1.15) | - | 0.09 | 0.94<br>(0.41-2.17) | 0.47<br>(0.11-1.98) | 0.16 | 1.58<br>(0.64-3.89) | 16.31<br>(1.96-135.84) | 0.05 | 0.58<br>(0.20-1.73) | 19.21<br>(0.00-283) |
| Central | 1945 | 0.54 | 1.02<br>(0.67-1.57) | - | 0.22 | **2.45**<br>(1.07-5.61)* | 0.84<br>(0.21-3.34) | 0.09 | 0.92<br>(0.33-2.56) | 8.43<br>(0.64-110.41) | 0.14 | 1.72<br>(0.59-4.99) | 0.00<br>(0.00-184) |
| Rift Valley | 3808 | 0.73 | 0.70<br>(0.49-1.02) | - | 0.17 | 0.94<br>(0.45-1.99) | 0.34<br>(0.11-1.03) | 0.23 | 1.26<br>(0.55-2.85) | 6.23<br>(0.71-54.95) | 0.17 | 1.10<br>(0.44-2.74) | 459 (0.00-1611) |
| Western | 1499 | 0.37 | 0.90<br>(0.57-1.43) | - | 0.10 | 1.34<br>(0.57-3.17) | 0.99<br>(0.32-3.07) | 0.16 | 2.21<br>(0.94-5.21) | **17.73**<br>(1.75-179.95)* | 0.18 | **2.98**<br>(1.15-7.70)* | 0.53<br>(0.00-413) |
| Nyanza | 1610 | 0.40 | 0.92<br>(0.62-1.37) | - | 0.15 | 2.04<br>(0.97-4.32) | 1.03<br>(0.25-4.23) | 0.13 | 1.64<br>(0.69-3.88) | **13.97**<br>**(1.12-**<br>**174.52)\*** | 0.08 | 1.19<br>(0.44-3.22) | 0.01<br>(0.00-370) |
| Nairobi | 1855 | 0.61 | 1.24<br>(0.71-2.19) | - | 0.17 | 1.95<br>(0.60-6.35) | 0.35<br>(0.10-1.25) | 0.36 | **4.18**<br>**(1.59-**<br>**10.97)\*\*** | **31.04**<br>**(3.22-**<br>**298.95)\*** | 0.28 | **3.77**<br>**(1.06-**<br>**13.39)\*** | 0.00<br>(0.00-370) |
| **Ethnicity** |  |  |  |  |  |  |  |  |  |  |  |  |  |
| Embu | 186 | 0.07 | 1.19<br>(0.60-2.35) | 1.31<br>(0.61-2.81) | 0.01 | 0.46<br>(0.10-2.05) | 0.00<br>(0.00-0.00) | 0.01 | 1.02<br>(0.20-5.21) | 0.53<br>(0.06-4.49) | 0.01 | 0.66<br>(0.16-2.70) | 0.36 (201-667) |
| Kalenjin | 1823 | 0.26 | **0.44**<br>**(0.28-**<br>**0.71)\*\*** | **0.53**<br>**(0.32-**<br>**0.88)\*\*** | 0.09 | **0.46**<br>**(0.23-**<br>**0.95)\*** | 1.32<br>(0.46-3.81) | 0.17 | 1.64<br>(0.64-4.21) | 2.69<br>(0.63-11.47) | 0.05 | **0.31**<br>**(0.13-**<br>**0.77)\*** | 578 (100-332) |
| Kamba | 1669 | 0.17 | **0.32**<br>**(0.16-**<br>**0.61)\*\*** | **0.30**<br>**(0.15-**<br>**0.56)\*\*** | 0.04 | **0.23**<br>**(0.09-**<br>**0.59)\*** | 0.43<br>(0.13-1.43) | 0.13 | 1.35<br>(0.48-3.79) | 0.85<br>(0.20-3.56) | 0.06 | 0.42<br>(0.15-1.22) | **54.88**<br>**(3.68-**<br>**819.16)\*** |
| Kikuyu | 2521 | 0.78 | 1 | 1 | 0.27 | 1 | 1 | 0.14 | 1 | 1 | 0.21 | 1 | 1 |
| Kisii | 901 | 0.15 | **0.53**<br>**(0.31-**<br>**0.91)\*\*** | **0.44**<br>**(0.23-**<br>**0.83)\*\*** | 0.06 | 0.59<br>(0.24-1.47) | 0.68<br>(0.17-2.74) | 0.005 | **0.09**<br>**(0.01-**<br>**0.60)\*** | **0.07**<br>**(0.01-**<br>**0.59)\*** | 0.07 | 0.91<br>(0.15-5.49) | 0.33 (410-274) |
| Luhya | 2424 | 0.53 | 0.69<br>(0.45-1.07) | 0.72<br>(0.46-1.12) | 0.11 | **0.44**<br>**(0.21-**<br>**0.93)\*** | 0.48<br>(0.14-1.59) | 0.21 | 1.49<br>(0.61-3.66) | 0.84<br>(0.20-3.57) | 0.20 | 0.98<br>(0.50-1.93) | **11.52**<br>**(1.73-**<br>**76.49)_\*** |
| Luo | 1591 | 0.56 | 1.15<br>(0.69-1.90) | 1.08<br>(0.66-1.79) | 0.17 | 0.99<br>(0.42-2.31) | 0.66<br>(0.11-4.12) | 0.17 | 1.90<br>(0.85-4.26) | 0.91<br>(0.22-3.68) | 0.17 | 1.30<br>(0.50-3.35) | 32.68<br>(0.04-271) |
| Maasai | 325 | 0.04 | 0.42<br>(0.14-1.24) | 0.61<br>(0.18-2.05) | 0.02 | 0.46<br>(0.10-2.07) | 2.01<br>(0.47-8.64) | 0.02 | 1.20<br>(0.31-4.65) | 1.70<br>(0.31-9.20) | 0.01 | 0.34<br>(0.07-1.56) | 5.08<br>(72.94-482.09) |
| Meru | 717 | 0.33 | 1.52<br>(0.85-2.72) | 1.40<br>(0.80-2.45) | 0.11 | 1.41<br>(0.39-5.15) | 1.56<br>(0.37-6.63) | 0.05 | 1.23<br>(0.39-3.92) | 0.70<br>(0.14-3.55) | 0.02 | 0.39<br>(0.12-1.29) | 101 (124-829) |
| Mijikenda/Swahili | 798 | 0.23 | 0.91<br>(0.57-1.47) | 1.39<br>(0.83-2.31) | 0.02 | **0.24**<br>**(0.07-**<br>**0.85)\*** | 0.24<br>(0.03-1.74) | 0.05 | 1.14<br>(0.37-3.56) | 11.04<br>(0.91-133.67) | 0.03 | 0.50<br>(0.13-1.98) | 185 (0.00-2.50) |
| Somali | 294 | 0.04 | 0.64<br>(0.31-1.31) | 0.53<br>(0.18-1.57) | 0.03 | 0.81<br>(0.39-1.68) | 0.50<br>(0.08-3.01) | 0.01 | 0.41<br>(0.12-1.46) | 0.59<br>(0.10-3.44) | 0.004 | 0.16<br>(0.02-1.20) | 538<br>(121.51-238) |
| Taita/Taveta | 136 | 0.06 | 1.05<br>(0.50-2.16) | 0.92<br>(0.38-2.21) | 0.01 | 0.50<br>(0.11-2.18) | 0.00<br>(0.00-0.00) | 0.08 | **10.79**<br>**(1.38-**<br>**84.17)\*** | **23.65**<br>**(1.19-**<br>**470.35)\*** | 0.004 | 0.34<br>(0.04-2.70) | 436.98<br>(534.08-356.23) |
| Other | 1068 | 0.26 | 0.78<br>(0.46-1.34) | 0.71<br>(0.37-1.34) | 0.07 | 0.61<br>(0.22-1.64) | 0.57<br>(0.14-2.30) | 0.14 | 2.35<br>(0.64-8.58) | 1.92<br>(0.38-9.76) | 0.11 | 1.28<br>(0.38-4.26) | 796.47<br>(974.54-6504.08) |
| **Education** |  |  |  |  |  |  |  |  |  |  |  |  |  |
| None/primary | 5660 | 1.30 | 1 | 1 | 0.30 | 1 | 1 | 0.47 | 1 | - | 0.24 | 1 | 1 |
| Secondary | 5851 | 1.10 | 0.82<br>(0.60-1.11) | 1.11<br>(0.77-1.62) | 0.29 | 0.94<br>(0.57-1.55) | 0.97<br>(0.54-1.75) | 0.59 | 1.22<br>(0.73-2.03) | - | 0.40 | **1.59**<br>**(0.93-**<br>**2.70)\*** | 3.80<br>(3.80-3.80) |
| Tertiary | 2943 | 1.09 | **1.66**<br>**(1.20-**<br>**2.29)\*\*** | **1.83**<br>**(1.19-**<br>**2.81)\*** | 0.42 | **2.74**<br>**(1.55-**<br>**4.82)\*\*** | **1.96**<br>**(1.03-**<br>**3.70)\*** | 0.14 | 0.57<br>(0.25-1.26) | - | 0.32 | **2.54**<br>**(1.27-**<br>**5.08)\*** | 0.00<br>(0.00-0.00) |
| **Religion** |  |  |  |  |  |  |  |  |  |  |  |  |  |
| Christians | 12361 | 3.04 | 1 | - | 0.86 | 1 | - | 1.09 | 1 | - | 0.82 | 1 | 1 |
| Muslims | 1042 | 0.30 | 1.17<br>(0.80-1.70) | - | 0.05 | 0.65<br>(0.36-1.21) | - | 0.04 | 0.47<br>(0.21-1.02) | - | 0.02 | 0.31<br>(0.12-0.76) | 0.00<br>(0.00-0.00) |
| Others | 1050 | 0.15 | 0.58<br>(0.31-1.08) | - | 0.09 | 1.26<br>(0.53-2.97) | - | 0.06 | 0.64<br>(0.21-1.95) | - | 0.12 | 1.77<br>(0.59-5.33) | 0.00<br>(0.00-0.00) |
| **Marital status** |  |  |  |  |  |  |  |  |  |  |  |  |  |
| Unmarried (single/<br>divorced/widowed/separated) | 7495 | 0.81 | 1 | 1 | 0.16 | 1 | 1 | 0.65 | 1 | - | 0.61 | 1 | - |
| Married/Cohabiting | 6958 | 2.68 | **3.70**<br>**(2.74-**<br>**4.99)\*** | 1.44<br>(0.96-2.15) | 0.84 | **5.84**<br>**(3.10-**<br>**10.98)\*\*** | 1.58<br>(0.81-3.10) | 0.54 | 0.90<br>(0.53-1.54) | - | 0.36 | 0.63<br>(0.34-1.15) | - |
| **Working status/occupation** |  |  |  |  |  |  |  |  |  |  |  |  |  |
| Not Working | 3157 | 0.29 | 1 | 1 | 0.04 | 1 | 1 | 0.31 | 1 | - | 0.16 | 1 | - |
| Working | 11296 | 3.20 | **3.12**<br>**(2.02-4.81)\*\*** | 1.13<br>(0.64-2.01) | 0.95 | **6.13**<br>**(2.33-16.13)\*\*** | 1.50<br>(0.39-5.81) | 0.88 | 0.79<br>(0.42-1.48) | - | 0.80 | 1.40<br>(0.57-3.45) | - |
| **Wealth index** |  |  |  |  |  |  |  |  |  |  |  |  |  |
| Poorest | 2175 | 0.37 | 1 | 1 | 0.04 | 1 | 1 | 0.19 | 1 | - | 0.13 | 1 | - |
| Poorer | 2762 | 0.48 | 1.02<br>(0.69-1.52) | 0.93<br>(0.59-1.48) | 0.09 | 1.61<br>(0.63-4.10) | 1.48<br>(0.51-4.31) | 0.25 | 1.05<br>(0.57-1.92) | - | 0.15 | 0.93<br>(0.50-1.76) | - |
| Middle | 2932 | 0.62 | 1.25<br>(0.86-1.83) | 1.04<br>(0.66-1.64) | 0.19 | **3.13**<br>**(1.32-7.41)\*\*** | **3.26**<br>**(1.16-9.20)\*** | 0.20 | 0.80<br>(0.43-1.47) | - | 0.13 | 0.73<br>(0.36-1.48) | - |
| Richer | 3504 | 0.83 | 1.40<br>(0.94-2.07) | 1.11<br>(0.67-1.83) | 0.22 | **3.08**<br>**(1.30-7.26)\*\*** | **3.50**<br>**(1.21-10.08)\*** | 0.29 | 0.96<br>(0.46-2.03) | - | 0.39 | 1.90<br>(0.83-4.38) | - |
| Richest | 3079 | 1.19 | **2.34**<br>**(1.55-3.53)\*\*** | 1.22<br>(0.73-2.06) | 0.45 | **7.26**<br>**(2.98-17.67)\*\*** | **7.55**<br>**(2.67-21.37)\*** | 0.25 | 0.93<br>(0.40-2.16) | - | 0.17 | 0.97<br>(0.37-2.56) | - |
| **Household number** |  |  |  |  |  |  |  |  |  |  |  |  |  |
| ≤4 | 717 | 0.14 | 1 | - | 0.09 | 1 | - | 0.03 | 1 | - | 0.02 | 1 | - |
| ≥5 | 13736 | 3.35 | 1.23<br>(0.68-2.23) | - | 0.90 | 0.49<br>(0.22-1.11) | - | 1.16 | 1.84<br>(0.61-5.53) | - | 0.94 | 1.99<br>(0.79-5.04) | - |
| **Health status** |  |  |  |  |  |  |  |  |  |  |  |  |  |
| Bad | 189 | 0.15 | 1 | 1 | 0.05 | 1 | 1 | 0.05 | 1 | 1 | 0.03 | 1 | 1 |
| Moderate | 2116 | 1.09 | 0.63<br>(0.36-1.09) | 0.53<br>(0.27-1.04) | 0.42 | 0.68<br>(0.27-1.68) | 0.51<br>(0.17-1.57) | 0.47 | 0.92<br>(0.32-2.63) | 0.78<br>(0.27-2.28) | 0.30 | 0.84<br>(0.30-2.35) | 0.03<br>(0.00-0.68) |
| Good | 12148 | 2.25 | **0.21**<br>**(0.13-0.36)\*\*** | **0.20**<br>**(0.10-0.38)\*\*** | 0.52 | **0.14**<br>**(0.06-0.33)\*\*** | **0.08**<br>**(0.03-0.23)\*\*** | 0.68 | **0.23**<br>**(0.08-0.63)\*\*** | **0.20**<br>**(0.07-0.58)\*\*** | 0.63 | **0.31**<br>**(0.12-0.81)\*\*** | 0.01<br>(0.00-0.32) |
| **Media access (composite)** |  |  |  |  |  |  |  |  |  |  |  |  |  |
| No | - | - | - | - | - | - | - | - | - | - | - | 1 | 1 |
| Yes | - | - | - | - | - | - | - | - | - | - | - | **0.33**<br>**(0.15-0.73)\*** | 65.08<br>(530.68-773.13) |
| **Newspaper** |  |  |  |  |  |  |  |  |  |  |  |  |  |
| No | 8753 | 1.86 | 1 | 1 | 0.51 | 1 | - | 0.83 | 1 | - | 0.52 | 1 | - |
| Yes | 5700 | 1.63 | **1.36**<br>**(1.04-1.78)\*** | 0.93<br>(0.68-1.28) | 0.48 | 1.46<br>(0.88-2.41) | - | 0.37 | 0.68<br>(0.41-1.14) | - | 0.44 | 1.28<br>(0.80-2.06) | - |
| **TV** |  |  |  |  |  |  | - |  |  |  |  |  |  |
| No | 2875 | 0.52 | 1 | 1 | 0.16 | 1 | - | 0.27 | 1 | - | 0.28 | 1 | - |
| Yes | 11578 | 2.97 | **1.43**<br>**(1.08-1.89)\*** | 1.17<br>(0.82-1.68) | 0.84 | 1.33<br>(0.78-2.25) | - | 0.93 | 0.87<br>(0.50-1.50) | - | 0.68 | 0.60<br>(0.35-1.03) | - |
| **Radio** |  |  |  |  |  |  | - |  |  | - |  |  |  |
| No | 1820 | 0.36 | 1 | - | 0.20 | 1 | - | 0.16 | 1 | - | 0.13 | 1 | - |
| Yes | 12633 | 3.13 | 1.26<br>(0.83-1.91) | - | 0.79 | 0.55<br>(0.29-1.07) | - | 1.03 | 0.94<br>(0.45-1.99) | - | 0.83 | 0.91<br>(0.44-1.90) | - |
| **Internet use** |  |  |  |  |  |  |  |  |  | - |  |  |  |
| No | 5957 | 1.20 | 1 | 1 | 0.30 | 1 | 1 | 0.51 | 1 | - | 0.39 | 1 | - |
| Yes | 8496 | 2.29 | **1.36**<br>**(1.03-1.79)\*** | 1.25<br>(0.85-1.84) | 0.70 | **1.63**<br>**(1.04-2.57)\*** | 1.00<br>(0.61-1.64) | 0.68 | 0.94<br>(0.59-1.52) | - | 0.57 | 0.99<br>(0.54-1.81) | - |
| **Lifestyle Behavioural factors** |  |  |  |  |  |  |  |  |  |  |  |  |  |
| **Tobacco use** |  |  |  |  |  |  |  |  |  |  |  |  | - |
| No | 12525 | 2.91 | 1 | - | 0.90 | 1 | - | 1.05 | 1 | - | 0.80 | 1 | - |
| Yes | 1928 | 0.58 | 1.30<br>(0.94-1.78) | - | 0.10 | 0.70<br>(0.32-1.50) | - | 0.14 | 0.90<br>(0.44-1.81) | - | 0.17 | 1.36<br>(0.70-2.64) | - |
| **Past smoking** |  |  |  | - |  |  | - |  |  |  |  |  |  |
| No | 288 | 1.10 | 1 | - | 0.29 | 1 | - | 0.40 | 1 | - | 0.10 | 1 | 1 |
| Yes | 206 | 1.45 | 1.86<br>(0.64-5.45) | - | 0.41 | 1.95<br>(0.15-25.48) | - | 0.68 | 2.39<br>(0.23-20.12) | - | 0.69 | **9.92**<br>**(1.40-70.03)\*** | 8.51<br>(0.86-84.00) |
| **Exercise Minutes per week** |  |  |  |  |  |  |  |  |  |  |  |  |  |
| Adequate ($\geq 150$minutes) | 10043 | 2.53 | 1 | 1 | 0.65 | 1 | 1 | 1.01 | 1 | - | 0.79 | 1 | - |
| Inadequate ($< 150$minutes) | 2426 | 0.87 | **1.44**<br>**(1.02-2.03)\*** | **1.51**<br>**(1.07-2.14)\*** | 0.36 | **2.28**<br>**(1.18-4.39)\*** | **1.90**<br>**(1.08-3.35)\*** | 0.19 | 0.79<br>(0.43-1.47) | - | 0.23 | 1.20<br>(0.53-2.75) | - |
| **Number of hours per day seated/sedentary** |  |  |  |  |  |  |  |  |  |  |  |  |  |
| Low risk ($\leq 8$hours)/non sedentary lie | 13692 | 3.26 | 1 | - | 0.85 | 1 | 1 | 1.15 | 1 | - | 0.94 | 1 | - |
| High-risk (>8hours)/sedentary life | 747 | 0.23 | 1.32<br>(0.67-2.60) | - | 0.15 | **3.41**<br>**(1.28-9.09)\*** | **2.90**<br>**(1.30-6.47)\*** | 0.05 | 0.75<br>(0.27-2.08) | - | 0.02 | 0.44<br>(0.17-1.15) | - |
| **Alcohol consumption** |  |  |  |  |  |  |  |  |  |  |  |  |  |
| No | 10608 | 2.21 | 1 | 1 | 0.70 | 1 | - | 0.90 | 1 | - | 0.65 | 1 | - |
| Yes | 3845 | 1.28 | **1.63**<br>**(1.23-2.16)\*\*** | 1.18<br>(0.85-1.63) | 0.30 | 1.18<br>(0.68-2.06) | - | 0.29 | 0.90<br>(0.52-1.55) | - | 0.31 | 1.32<br>(0.67-2.61) | - |

**Table 3. (continued)**
|  |  | **Depression** |  |  | **Anxiety** |  |  | **Arthritis** |  |  | **Atleast1NCD** |  |  |
| --- | --- | --- | --- | --- | --- | --- | --- | --- | --- | --- | --- | --- | --- |
| Variable | n | Yes,<br>% | uORs<br>(95CI) | aORs<br>(95CI) | Yes,<br>% | uORs<br>(95CI) | aORs<br>(95CI) | Yes,<br>% | uORs<br>(95CI) | aORs<br>(95CI) | Yes,<br>% | uORs<br>(95CI) | aORs (95CI) |
| **Age (years)** |  |  |  |  |  |  |  |  |  |  |  |  |  |
| 15-24 | 5579 | 0.24 | 1 | 1 | 0.51 | 1 | - | 0.24 | 1 | 1 | 2.14 | 1 | 1 |
| 25-34 | 4055 | 0.73 | **4.31**<br>**(2.76-6.73)\*\*** | **4.08**<br>**(2.40-6.93)\*\*** | 0.44 | 1.20<br>(0.65-2.19) | - | 0.19 | 1.07<br>(0.62-1.84) | 0.78<br>(0.39-1.56) | 1.98 | **0.46 (0.40-0.51)\*** | 1.10 (0.80-1.51) |
| 35-59 | 4819 | 1.23 | **6.15**<br>**(4.03-9.38)\*\*** | **5.61**<br>**(3.15-9.97)\*\*** | 0.62 | 1.44<br>(0.82-2.53) | - | 0.93 | **4.56**<br>**(2.87-7.24)\*\*** | **2.62**<br>**(1.33-5.13)\*\*** | 5.29 | **0.82 (0.72-0.94)\*** | **2.94 (2.17-3.97)\*** |
| **Residence** |  |  |  |  |  |  |  |  |  |  |  |  |  |
| Urban | 5624 | 0.76 | 0.83<br>(0.59-1.16) | - | 0.78 | **1.55**<br>**(1.01-2.38)\*\*\*** | 1.10<br>(0.69-1.76) | 0.23 | **0.32**<br>**(0.19-0.55)\*\*** | **0.37**<br>**(0.21-0.61)\*\*** | 3.78 | **1.30 (1.13-1.50)\*\*** | **0.76 (0.59-0.97)\*** |
| Rural | 8829 | 1.44 | 1 | 1 | 0.79 | 1 | 1 | 1.13 | 1 | 1 | 5.64 | 1 | 1 |
| **Region/Province** |  |  |  |  |  |  |  |  |  |  |  |  |  |
| Coast | 1341 | 0.14 | 1 | 1 | 0.05 | 1 | 1 | 0.10 | 1 | 1 | 0.68 | 1 | 1 |
| Northeastern | 273 | 0.001 | **0.04**<br>**(0.01-0.32)\*\*** | 0.16<br>(0.02-1.30) | 0.003 | 0.33<br>(0.07-1.48) | 0.69<br>(0.00.13-3.68) | 0.00 | 0.00<br>(0.00-0.00) | 0.00<br>(0.00-0.00) | 0.06 | **0.51 (0.37-0.73)\*** | 0.95 (0.31-2.93) |
| Eastern | 2123 | 0.17 | 0.73<br>(0.40-1.31) | 0.77<br>(0.32-1.86) | 0.11 | 1.52<br>(0.76-3.04) | 0.56<br>(0.22-1.40) | 0.48 | **3.04**<br>**(1.53-6.05)\*** | 0.85<br>(0.23-3.14) | 1.29 | **1.55 (1.31-1.83)\*** | 1.25 (0.66-2.37) |
| Central | 1945 | 0.21 | 1.01<br>(0.52-1.97) | 1.08<br>(0.35-3.34) | 0.22 | **3.30**<br>**(1.51-7.24)\*\*** | 0.52<br>(0.17-1.57) | 0.23 | 1.56<br>(0.80-3.06) | 1.07<br>(0.26-4.29) | 1.31 | **1.56 (1.26-1.94)\*** | 1.15 (0.54-2.43) |
| Rift Valley | 3808 | 0.88 | **2.20**<br>**(1.35-3.60)\*\*** | **2.74**<br>**(1.14-6.58)\*\*** | 0.41 | **3.06**<br>**(1.64-5.72)\*\*** | **0.30**<br>**(0.12-0.73)\*\*** | 0.32 | 1.12<br>(0.57-2.18) | 1.22<br>(0.31-4.82) | 2.45 | 0.91 (0.78-1.06) | 1.54 (0.80-2.95) |
| Western | 1499 | 0.33 | **2.08**<br>**(1.15-3.79)\*\*** | **2.92**<br>**(1.13-7.53)\*\*** | 0.30 | **5.72**<br>**(3.00-10.90)\*\*** | **0.11**<br>**(0.04-0.30)\*\*** | 0.10 | 0.86<br>(0.38-1.98) | 0.62<br>(0.14-2.65) | 1.08 | 1.08 (0.88-1.32) | **2.23 (1.13-4.38)\*\*** |
| Nyanza | 1610 | 0.18 | 1.07<br>(0.60-1.91) | 1.33<br>(0.46-3.83) | 0.13 | **2.21**<br>**(1.08-4.55)\*\*** | **0.20**<br>**(0.06-0.71)\*\*** | 0.08 | 0.69<br>(0.31-1.54) | 0.21<br>(0.03-1.35) | 0.87 | **0.77 (0.65-0.91)\*** | 1.20 (0.53-2.74) |
| Nairobi | 1855 | 0.28 | 1.43<br>(0.64-3.21) | 1.73<br>(0.60-4.93) | 0.35 | **5.48**<br>**(2.25-13.34)\*\*** | **0.23**<br>**(0.08-0.63)\*\*\*** | 0.05 | 0.32<br>(0.04-2.42) | 0.67<br>(0.07-6.27) | 1.68 | **2.16 (1.53-3.07)\*** | **2.39 (1.18-4.86)\*\*** |
| **Ethnicity** |  |  |  |  |  |  |  |  |  |  |  |  |  |
| Embu | 186 | 0.01 | 0.49<br>(0.15-1.59) | 0.61<br>(0.16-2.29) | 0.01 | 0.41<br>(0.13-1.36) | 1.78<br>(0.45-7.05) | 0.03 | 1.26<br>(0.49-3.27) | 1.06<br>(0.32-3.53) | 0.13 | 1.11 (0.76-1.63) | 0.65 (0.34-1.25) |
| Kalenjin | 1823 | 0.46 | **1.70**<br>**(1.09-2.66)\*\*** | 1.09<br>(0.61-1.96) | 0.20 | 0.57<br>(0.31-1.07) | 1.80<br>(0.86-3.75) | 0.10 | **0.48**<br>**(0.26-0.92)\*** | **0.34**<br>**(0.14-0.82)\*** | 1.18 | 0.56 (0.46-0.68)** | 0.74 (0.52-1.06) |
| Kamba | 1669 | 0.19 | 0.75<br>(0.34-1.61) | 1.03<br>(0.32-3.29) | 0.12 | 0.37<br>(0.13-1.07) | 2.29<br>(0.53-9.89) | 0.06 | **0.29**<br>**(0.12-0.67)\*** | **0.25**<br>**(0.07-0.85)\*** | 0.54 | 0.98 (0.80-1.19) | **0.36 (0.21-0.63)\*\*** |
| Kikuyu | 2521 | 0.38 | 1 | 1 | 0.48 | 1 | 1 | 0.29 | 1 | 1 | 2.09 | 1 | 1 |
| Kisii | 901 | 0.10 | 0.74<br>(0.21-2.59) | 0.77<br>(0.18-3.31) | 0.02 | **0.10**<br>**(0.03-0.31)** | **12.90**<br>**(3.36-49.46)\*** | 0.02 | **0.16**<br>**(0.04-0.66)\*** | 0.22<br>(0.03-1.70) | 0.37 | **0.71 (0.54-0.92)\*\*** | 0.48 (0.21-1.10) |
| Luhya | 2424 | 0.47 | 1.29<br>(0.80-2.10) | 0.79<br>(0.39-1.60) | 0.37 | 0.81<br>(0.46-1.43) | **2.43**<br>**(1.09-5.42)\*** | 0.14 | **0.51**<br>**(0.28-0.93)\*\*** | 0.64<br>(0.28-1.44) | 1.51 | **0.73 (0.60-0.89)\*\*** | **0.52 (0.35-0.79)\*\*** |
| Luo | 1591 | 0.24 | 0.99<br>(0.54-1.81) | 0.89<br>(0.35-2.25) | 0.19 | 0.62<br>(0.27-1.43) | 3.09<br>(0.77-12.44) | 0.12 | 0.67<br>(0.28-1.60) | 1.86<br>(0.50-6.94) | 1.23 | **0.58 (0.42-0.81)\*\*** | 0.73 (0.40-1.35) |
| Maasai | 325 | 0.07 | 1.39<br>(0.70-2.78) | 1.09<br>(0.49-2.40) | 0.02 | 0.32<br>(0.05-1.93) | 2.80<br>(0.44-17.73) | 0.05 | 1.44<br>(0.52-4.00) | 0.70<br>(0.23-2.07) | 0.17 | **0.48 (0.35-**<br>**0.66)\*\*** | 0.66 (0.35-1.24) |
| Meru | 717 | 0.08 | 0.75<br>(0.37-1.52) | 0.94<br>(0.37-2.39) | 0.07 | 0.48<br>(0.22-1.05) | 1.45<br>(0.55-3.82) | 0.41 | **5.41**<br>**(3.16-**<br>**9.24)\*** | **3.59**<br>**(1.72-**<br>**7.50)\*** | 0.87 | **1.44 (1.11-**<br>**1.87)\*\*** | 1.44 (0.90-2.29) |
| Mijikenda/Swahili | 798 | 0.10 | 0.80<br>(0.42-1.53) | 0.77<br>(0.27-2.21) | 0.04 | **0.24**<br>**(0.11-**<br>**0.53)\*** | 1.26<br>(0.42-3.80) | 0.07 | 0.80<br>(0.36-1.75) | 0.59<br>(0.12-2.81) | 0.43 | **0.59 (0.47-**<br>**0.74)\*\*** | 0.83 (0.39-1.79) |
| Taita/Taveta | 136 | 0.01 | 0.69<br>(0.19-2.49) | 0.79<br>(0.18-3.53) | 0.001 | 0.00<br>(0.00-0.00) | 260.09<br>(944.07-72052.29) | 0.01 | 0.63<br>(0.14-2.80) | 0.45<br>(0.04-4.61) | 0.14 | 0.69 (0.45-1.07) | 1.53 (0.31-7.50) |
| Somali | 294 | 0.003 | **0.08**<br>**(0.02-**<br>**0.27)\*\*** | **0.10**<br>**(0.02-**<br>**0.47)\*** | 0.005 | **0.08**<br>**(0.02-**<br>**0.28)\*** | **8.96**<br>**(1.84-**<br>**43.64)** | 0.00 | 0.00<br>(0.00-0.00) | 0.00<br>(0.00-0.00) | 0.09 | **0.35 (0.26-**<br>**0.47)\*\*** | 0.39 (0.12-1.26) |
| Other | 1068 | 0.11 | 0.68<br>(0.35-1.32) | **0.46**<br>**(0.22-**<br>**0.96)\*** | 0.05 | **0.25**<br>**(0.09-**<br>**0.69)\*** | **5.24**<br>**(1.61-**<br>**17.04)\*** | 0.06 | **0.45**<br>**(0.23-**<br>**0.87)\*** | 0.42<br>(0.18-1.00) | 0.66 | **0.72 (0.58-**<br>**0.90)\*\*** | 0.63 (0.37-1.06) |
| **Education** |  |  |  |  |  |  |  |  |  |  |  |  |  |
| None/primary | 5660 | 1.03 | 1 | 1 | 0.47 | 1 | 1 | 0.82 | 1 | 1 | 3.71 | 1 | 1 |
| Secondary | 5851 | 0.60 | **0.56**<br>**(0.41-**<br>**0.74)\*\*** | 0.91<br>(0.64-1.30) | 0.56 | 1.17<br>(0.70-1.96) | 0.94<br>(0.60-1.49) | 0.30 | **0.35**<br>**(0.24-**<br>**0.52)\*\*** | **0.54**<br>**(0.34-**<br>**0.86)\*** | 3.13 | **0.80 (0.73-**<br>**0.89)\*\*** | 1.03 (0.85-1.25) |
| Tertiary | 2943 | 0.58 | 1.08<br>(0.71-1.64) | 1.58<br>(0.99-2.51) | 0.54 | **2.23**<br>**(1.37-**<br>**3.64)\*\*** | 0.64<br>(0.34-1.19) | 0.24 | **0.56**<br>**(0.32-**<br>**0.99)\*\*** | 1.32<br>(0.71-2.45) | 2.57 | 1.15 (0.96-1.37) | **1.58 (1.20-**<br>**2.08)\*\*** |
| **Religion** |  |  |  |  |  |  |  |  |  |  |  |  |  |
| Christians | 12361 | 1.81 | 1 | 1 | 1.34 | 1 | - | 1.04 | 1 | 1 | 7.89 | 1 | 1 |
| Muslims | 1042 | 0.16 | 1.05<br>(0.63-1.74) | **3.16**<br>**(1.66-**<br>**6.04)\*\*** | 0.08 | 0.69<br>(0.38-1.26) | - | 0.06 | 0.65<br>(0.29-1.46) | 1.75<br>(0.68-4.51) | 0.59 | **0.70 (0.59-**<br>**0.84)\*\*** | **1.53 (1.05-**<br>**2.23)\*** |
| Others | 1050 | 0.23 | **1.52**<br>**(1.03-**<br>**2.25)\*\*\*** | **1.52**<br>**(1.01-**<br>**2.26)\*\*** | 0.15 | 1.31<br>(0.53-3.22) | - | 0.26 | **3.03**<br>**(1.67-**<br>**5.50)\*\*** | **2.12**<br>**(1.17-**<br>**3.84)\*** | 0.94 | **1.34 (1.09-**<br>**1.64)\*\*** | **1.55 (1.03-**<br>**2.31)\*** |
| **Marital status** |  |  |  |  |  |  |  |  |  |  |  |  |  |
| Unmarried (single/<br>divorced/widowed/separated) | 7495 | 0.80 | 1 | 1 | 0.85 | 1 | - | 0.49 | 1 | 1 | 3.62 | **1** | 1 |
| Married/Cohabiting | 6958 | 1.40 | **1.90**<br>**(1.39-2.61)\*\*** | 0.77<br>(0.51-1.16) | 0.71 | 0.90<br>(0.58-1.40) | - | 0.87 | **1.93**<br>**(1.25-2.99)\*\*** | 1.08<br>(0.63-1.84) | 5.80 | **1.27 (1.14-1.41)\*\*** | 0.98 (0.79-1.22) |
| **Working status/occupation** |  |  |  |  |  |  |  |  |  |  |  |  |  |
| Not Working | 3157 | 0.19 | 1 | 1 | 0.34 | ** | - | 0.22 | 1 | - | 1.40 | 1 | 1 |
| Working | 11296 | 2.01 | **2.96**<br>**(1.87-4.69)\*\*** | 1.30<br>(0.78-2.16) | 1.23 | 1.00<br>(0.50-2.00) | - | 1.14 | 1.43<br>(0.92-2.22) | - | 8.02 | **1.47 (1.29-1.67)\*\*** | 1.00 (0.73-1.37) |
| **Wealth index** |  |  |  |  |  |  |  |  |  |  |  |  |  |
| Poorest | 2175 | 0.34 | 1 | - | 0.18 | 1 | 1 | 0.22 | 1 | - | 1.22 | 1 | 1 |
| Poorer | 2762 | 0.49 | 1.15<br>(0.76-1.73) | - | 0.24 | 1.01<br>(0.56-1.85) | 1.19<br>(0.64-2.19) | 0.43 | 1.59<br>(0.88-2.88) | - | 1.69 | 0.90 (0.79-1.02) | 1.06 (0.82-1.36) |
| Middle | 2932 | 0.45 | 0.98<br>(0.65-1.49) | - | 0.24 | 0.95<br>(0.54-1.68) | 1.21<br>(0.67-2.21) | 0.29 | 0.99<br>(0.56-1.76) | - | 1.66 | **0.81 (0.71-0.92)\*\*** | 1.05 (0.81-1.35) |
| Richer | 3504 | 0.52 | 0.96<br>(0.60-1.54) | - | 0.25 | 0.85<br>(0.46-1.59) | 1.35<br>(0.66-2.75) | 0.25 | 0.70<br>(0.39-1.26) | - | 2.25 | 0.90 (0.77-1.05) | 1.28 (0.96-1.71) |
| Richest | 3079 | 0.40 | 0.82<br>(0.48-1.41) | - | 0.66 | **2.60**<br>**(1.46-4.64)\*\*** | **0.50**<br>**(0.26-0.94)\*** | 0.18 | 0.58<br>(0.27-1.27) | - | 2.60 | **1.41 (1.17-1.69)\*\*** | **1.59 (1.05-2.41)\*** |
| **Household number** |  |  |  |  |  |  |  |  |  |  |  |  |  |
| ≤4 | 717 | 0.08 | 1 | - | 0.09 | 1 | - | 0.06 | 1 | - | 0.38 | 1 | - |
| ≥5 | 13736 | 2.12 | 1.32<br>(0.74-2.34) | - | 1.48 | 0.85<br>(0.34-2.15) | - | 1.30 | 1.06<br>(0.43-2.59) | - | 9.04 | 0.95 (0.75-1.20) | - |
| **Health status** |  |  |  |  |  |  |  |  |  |  |  |  |  |
| Bad | 189 | 0.14 | 1 | 1 | 0.10 | 1 | 1 | 0.10 | 1 | 1 | 0.37 | 1 | 1 |
| Moderate | 2116 | 0.65 | **0.38**<br>**(0.21-0.70)\*\*** | 0.00<br>(0.00-0.00) | 0.36 | **0.29**<br>**(0.14-0.62)\*\*** | **3.83**<br>**(1.72-8.54)\*\*** | 0.47 | **0.41**<br>**(0.21-0.80)\*\*** | 0.46<br>(0.19-1.15) | 2.78 | **0.65 (0.45-0.94)\*** | **0.22 (0.15-0.34)\*** |
| Good | 12148 | 1.41 | **0.14**<br>**(0.08-0.25)\*\*** | 0.00<br>(0.00-0.00) | 1.10 | **0.15**<br>**(0.08-0.31)\*\*** | **7.38**<br>**(3.61-15.08)\*\*** | 0.80 | **0.12**<br>**(0.06-0.23)\*\*** | **0.17**<br>**(0.07-0.39)\*\*** | 6.27 | **0.61 (0.43-0.87)\*** | **0.62 (0.41-0.96)\*** |
| **Newspaper** |  |  |  |  |  |  |  |  |  |  |  |  |  |
| No | 8753 | 1.41 | 1 | - | 0.85 | 1 | - | 0.93 | 1 | - | 5.49 | 1 | 1 |
| Yes | 5700 | 0.79 | 0.86<br>(0.62-1.18) | - | 0.71 | 1.29<br>(0.84-1.99) | - | 0.43 | 0.71<br>(0.48-1.07) | - | 3.93 | **1.13 (1.02-**<br>**1.25)\*** | 0.95 (0.78-1.16) |
| **TV** |  |  |  |  |  |  |  |  |  |  |  |  |  |
| No | 2875 | 0.52 | 1 | - | 0.26 | 1 | - | 0.28 | 1 | - | 1.79 | 1 | - |
| Yes | 11578 | 1.68 | 0.79<br>(0.55-1.14) | - | 1.31 | 1.25<br>(0.76-2.03) | - | 1.08 | 0.95<br>(0.62-1.47) | - | 7.63 | 1.07 (0.94-1.20) | - |
| **Radio** |  |  |  |  |  |  |  |  |  |  |  |  | - |
| No | 1820 | 0.26 | 1 | - | 0.33 | 1 | 1 | 0.17 | 1 | - | 1.28 | 1 | - |
| Yes | 12633 | 1.94 | 1.07<br>(0.67-1.71) | - | 1.23 | **0.53**<br>**(0.29-**<br>**0.95)\*** | **1.79**<br>**(1.02-**<br>**3.13)** | 1.19 | 1.01<br>(0.59-1.73) | - | 8.14 | 1.10 (0.93-1.30) | - |
| **Internet use** |  |  |  |  |  |  |  |  |  |  |  |  | - |
| No | 5957 | 1.09 | 1 | 1 | 0.49 | 1 | 1 | 0.79 | 1 | 1 | 3.81 | 1 | - |
| Yes | 8496 | 1.12 | **0.72**<br>**(0.54-**<br>**0.95)\*** | 0.00<br>(0.00-0.00) | 1.08 | **1.54**<br>**(1.05-**<br>**2.27)\*** | 0.90<br>(0.58-1.40) | 0.57 | **0.51**<br>**(0.35-**<br>**0.74)\*\*** | 1.00<br>(0.65-1.53) | 5.61 | 0.97 (0.87-1.08) | - |
| **Lifestyle Behavioural factors** |  |  |  |  |  |  |  |  |  |  |  |  |  |
| **Tobacco use** |  |  |  |  |  |  |  |  |  |  |  |  | - |
| No | 12525 | 1.71 | 1 | 1 | 1.34 | 1 | - | 0.94 | 1 | 1 | 7.70 | 1 | - |
| Yes | 1928 | 0.50 | **1.94**<br>**(1.39-**<br>**2.73)\*\*** | 1.13<br>(0.73-1.75) | 0.23 | 1.09<br>(0.70-1.71) | - | 0.42 | **2.96**<br>**(2.01-**<br>**4.36)\*\*** | 1.06<br>(0.68-1.64) | 1.72 | 5.36 (0.00-2.16) | - |
| **Smokeless tobacco** |  |  |  |  |  |  |  |  |  |  |  |  |  |
| No | - | - | - | - | - | - | - | - | - | - | 9.11 | 1 | - |
| Yes | - | - | - | - | - | - | - | - | - | - | 0.31 | 5.39<br>(5.39-5.39) | - |
| **Smoking tobacco** |  |  |  |  |  |  |  |  |  |  |  |  | - |
| No | - | - | - | - | - | - | - | - | - | - | 7.85 | 1 | - |
| Yes | - | - | - | - | - | - | - | - | - | - | 1.57 | 5.16 (0.00-1.34) | - |
| **Past smoking** |  |  |  |  |  |  |  |  |  |  |  |  |  |
| No | 288 | 2.68 | 1 | - | 1.27 | 1 | - | 1.40 | 1 | - | 6.30 | - | - |
| Yes | 206 | 1.78 | 0.92<br>(0.27-3.17) | - | 0.00 | 0.00<br>(0.00-0.00) | - | 2.54 | 2.63<br>(0.81-8.59) | - | 6.01 | - | - |
| **Exercise Minutes per week** |  |  |  |  |  |  | - |  |  |  |  |  | - |
| Adequate ($\geq 150$minutes) | 10043 | 1.97 | 1 | - | 1.34 | 1 | - | 1.27 | 1 | 1 | 7.61 | 1 | - |
| Inadequate ($< 150$minutes) | 2426 | 0.37 | 0.78<br>(0.50-1.21) | - | 0.26 | 0.79<br>(0.42-1.50) | - | 0.13 | **0.41**<br>**(0.22-**<br>**0.78)\*** | 0.51<br>(0.24-1.08) | 1.88 | 5.97 (5.97-59.86) | - |
| **Number of hours per day seated/sedentary** |  |  |  | - |  |  |  |  |  |  |  |  |  |
| Low risk ($\leq 8$hours)/ non sedentary | 13692 | 2.10 | 1 | - | 1.49 | 1 | - | 1.28 | 1 | - | 8.86 | 1 | - |
| High risk<br>($> 8$hours)/sedentary life | 747 | 0.10 | 0.90<br>(0.49-1.64) | - | 0.06 | 0.70<br>(0.28-1.73) | - | 0.08 | 1.18<br>(0.46-3.05) | - | 0.55 | 15.13<br>(0.00-3.26) | - |
| **Alcohol consumption** |  |  |  |  |  |  |  |  |  | - |  |  |  |
| No | 10608 | 1.36 | 1 | 1 | 1.06 | 1 | - | 0.94 | 1 | - | 6.33 | 1 | - |
| Yes | 3845 | 0.84 | **1.73**<br>**(1.28-**<br>**2.34)\*\*** | 1.17<br>(0.84-1.65) | 0.51 | 1.34<br>(0.85-2.13) | - | 0.42 | 1.24<br>(0.82-1.88) | - | 3.09 | 245.35<br>(225.83-265.01) | - |

Note. OR=unadjusted odd ratios, aOR=adjusted odds ratio\* = significant at $p < 0.05$ , \*\* = significant at $p < 0.001$ , \*\*\* = significant at $p < 0.1$ , – = not evaluated in that model, n=row total, %= percentage of the total sample size when all the row totals are added up.

**Table 4.** Factors associated with having multiple NCDs (multi-morbidity) among men in Kenya

| Variable | Multiple NCDs ($\geq 2$)/multi-morbidity | | | |
| --- | --- | --- | --- | --- |
|  | n | Yes,<br>% | uORs (95CI) | aORs (95CI) |
| **Age(years)** |  |  |  |  |
| 15-24 | 5579 | 0.19 | 1 | 1 |
| 25-34 | 4055 | 0.47 | **3.54 (1.55-8.06)\*** | 2.61 (0.97-7.01) |
| 35-59 | 4819 | 1.27 | **8.17 (3.72-17.95)\*** | **6.95 (2.66-18.19)\*\*** |
| **Residence** |  |  |  |  |
| Urban | 5624 | 0.79 | 1.10 (0.75-1.62) | - |
| Rural | 8829 | 1.13 | 1 | - |
| **Region/Province** |  |  |  |  |
| Coast | 1341 | 0.14 | 1 | 1 |
| Northeastern | 273 | 0.01 | 0.32 (0.10-1.00) | 2.35 (0.75-7.34) |
| Eastern | 2123 | 0.18 | 0.86 (0.47-1.58) | 0.70 (0.29-1.71) |
| Central | 1945 | 0.26 | 1.35 (0.72-2.53) | 1.56 (0.57-4.29) |
| RiftValley | 3808 | 0.40 | 1.05 (0.63-1.74) | 1.79 (0.83-3.86) |
| Western | 1499 | 0.29 | **1.96 (1.12-3.43) \*\*** | **3.93 (1.72-8.96)\*** |
| Nyanza | 1610 | 0.23 | 1.45 (0.85-2.48) | 1.79 (0.69-4.61) |
| Nairobi | 1855 | 0.41 | **2.23 (1.08-4.60) \*\*** | 2.35 (0.99-5.59) |
| **Ethnicity** |  |  |  |  |
| Embu | 186 | 0.02 | 0.77 (0.27-2.16) | 0.95 (0.27-3.37) |
| Kalenjin | 1823 | 0.14 | **0.54 (0.30-0.97) \*** | 0.63 (0.29-1.38) |
| Kamba | 1669 | 0.19 | 0.76 (0.35-1.64) | 1.16 (0.39-3.45) |
| Kikuyu | 2521 | 0.37 | 1 | 1 |
| Kisii | 901 | 0.04 | **0.31 (0.12-0.78) \*** | **0.33 (0.11-0.96)\*\*** |
| Luhya | 2424 | 0.35 | 1.00 (0.60-1.65) | 0.62 (0.29-1.32) |
| Luo | 1591 | 0.36 | 1.56 (0.85-2.85) | 1.14 (0.38-3.44) |
| Maasai | 325 | 0.04 | 0.93 (0.40-2.18) | 1.48 (0.54-4.05) |
| Meru | 717 | 0.16 | 1.60 (0.63-4.07) | 2.70 (0.91-7.98) |
| Mijikenda/Swahili | 798 | 0.10 | 0.84 (0.43-1.62) | 2.12 (0.73-6.15) |
| Taita/Taveta | 136 | 0.01 | 0.69 (0.20-2.31) | 0.96 (0.24-3.81) |
| Somali | 294 | 0.01 | **0.20 (0.07-0.63)\*** | 0.27 (0.07-1.01) |
| Other | 1068 | 0.14 | 0.87 (0.30-2.48) | 0.96 (0.25-3.73) |
| **Education** |  |  |  |  |
| None/primary | 5660 | 0.70 | 1 | 1 |
| Secondary | 5851 | 0.60 | 0.83 (0.56-1.23) | 1.28 (0.87-1.89) |
| Tertiary | 2943 | 0.63 | **1.74 (1.12-2.70)\*** | **2.03 (1.28-3.22)\*** |
| **Religion** |  |  |  |  |
| Christians | 12361 | 1.73 | 1 | - |
| Muslims | 1042 | 0.11 | 0.73 (0.41-1.29) | - |
| Others | 1050 | 0.09 | 0.63 (0.34-1.18) | - |
| **Marital status** |  |  |  |  |
| Unmarried(single/divorced/widowed/separated) | 7495 | 0.61 | 1 | 1 |
| Married/Cohabiting | 6958 | 1.31 | **2.34 (1.57-3.47)\*\*** | 0.78 (0.47-1.30) |
| **Working status/occupation** |  |  |  |  |
| Not Working | 3157 | 0.14 | 1 | 1 |
| Working | 11296 | 1.79 | **3.62 (1.97-6.65)\*\*** | 1.64 (0.72-3.76) |
| **Wealth index** |  |  |  |  |
| Poorest | 2175 | 0.22 | 1 | 1 |
| Poorer | 2762 | 0.35 | 1.27 (0.76-2.13) | 1.17 (0.68-2.00) |
| Middle | 2932 | 0.34 | 1.16 (0.68-1.96) | 1.18 (0.67-2.06) |
| Richer | 3504 | 0.40 | 1.13 (0.64-2.02) | 1.07 (0.55-2.08) |
| Richest | 3079 | 0.61 | **1.97 (1.11-3.50)\*\*\*** | 1.55 (0.83-2.91) |
| **Household number** |  |  |  |  |
| ≤4 | 717 | 0.1 | 1 | - |
| ≥5 | 13736 | 1.8 | 0.86 (0.421-1.74) | - |
| **Health status** |  |  |  |  |
| Bad | 189 | 0.16 | 1 | 1 |
| Moderate | 2116 | 0.76 | **0.09 (0.05-0.16)\*** | **0.37 (0.2-0.69)\*\*** |
| Good | 12148 | 1.01 | **0.41 (0.22-0.76)\*** | **0.10 (0.05-0.18)\*\*** |
| **Newspaper** |  |  |  |  |
| No | 8753 | 1.16 | 1 | - |
| Yes | 5700 | 0.77 | 1.02 (0.70-1.49) | - |
| **TV** |  |  |  |  |
| No | 2875 | 0.38 | 1 | - |
| Yes | 11578 | 1.55 | 1.01 (0.68-1.51) | - |
| **Radio** |  |  |  |  |
| No | 1820 | 0.23 | 1 | - |
| Yes | 12633 | 1.69 | 1.05 (0.61-1.80) | - |
| **Internet use** |  |  |  |  |
| No | 5957 | 0.69 | 1 | - |
| Yes | 8496 | 1.24 | 1.27 (0.92-1.76) | - |
| **Smokeless tobacco** |  |  |  |  |
| No | 14171 | 1.89 | 1 | - |
| Yes | 281 | 0.04 | 0.99 (0.36-2.77) | - |
| **Smokingtobacco** |  |  |  |  |
| No | 12711 | 1.57 | 1 | 1 |
| Yes | 1741 | 0.36 | **1.71 (1.16-2.50)\*** | 1.07 (0.66-1.74) |
| **Past smoking** |  |  |  |  |
| No | 288 | 0.95 | 1 | - |
| Yes | 206 | 1.54 | 2.32 (0.61-8.85) | - |
| **Lifestyle Behavioural factors** |  |  |  |  |
| **Exercise Minutes per week** |  |  |  |  |
| Adequate(≥150minutes) | 10043 | 1.58 | 1 | - |
| Inadequate(<150minutes) | 2426 | 0.47 | 1.23 (0.73-2.07) | - |
| **Number of hours per days seated/sedentary** |  |  |  |  |
| Low risk(≤8hours)/non sedentary | 13692 | 1.78 | 1 | - |
| High risk(>8hours)/sedentary life | 747 | 0.15 | 1.53 (0.60-3.91) | - |
| **Alcohol consumption** |  |  |  |  |
| No | 10608 | 1.20 | 1 | 1 |
| Yes | 3845 | 0.73 | **1.69 (1.18-2.43)\*\*** | 1.05 (0.68-1.62) |

Note. OR=unadjusted odd ratios, aOR=adjusted odds ratio\* = significant at p<0.05, \*\* = significant at p<0.001, \*\*\* = significant at p<0.1, – = not evaluated in that model, n=row total, %= percentage of the total sample size when all the row totals are added up.

The findings of this study also indicated men from other regions of Kenya when compared to those from the coastal region, were more likely to have a heart disease (e.g., from the Northeastern (aOR 6.51 (95%CI: 1.11 - 38.21), western (aOR 17.73 (95%CI:1.75-179.95), Nyanza (aOR 13.97 (95%CI:1.12-174.52) and Nairobi (aOR 31.04 (95%CI: 3.22-298.95), experience depression (e.g., Rift valley (aOR 2.74 (95%CI:1.14-6.58) and western (aOR 2.92 (95%CI: 1.13-7.53), have at least one NCD (e.g., western (aOR 2.23 (95%CI:1.13-4.38), and multiple NCDs (e.g., western province =(aOR 3.93 (95%CI:1.72-8.96). However, those from the Rift Valley (aOR 0.30 (95%CI:0.12 - 0.73), Western (aOR 0.11 (95%CI:0.04-0.30), Nyanza (aOR 0.20 (95%CI:0.06 -0.71), and Nairobi (aOR 0.23 (95%CI:0.08-0.63) provinces of Kenya when compared with those from the coastal region, were less likely to experience anxiety.

We found that experiencing NCDs also varied across tribes. Compared with the Kikuyu tribe, other tribes of Kenya had higher odds of having lung disease (e.g., Luhya =aOR 11.52 (95%CI:1.73-76.49), heart disease (e.g., Taita/Taveta =aOR 23.65 95%CI: 1.19-470.35), experiencing anxiety (e.g., Luhya (aOR 2.43 (95%CI:1.09-5.42), and having arthritis (e.g., Meru tribe = (aOR 3.59 (95%CI:1.72-7.50). However, some ethnic tribes had lower odds of experiencing heart disease (e.g., Kisii tribe =aOR 0.07 95%CI: 0.01-0.59), depression (e.g., Somali (aOR 0.10 (95%CI:0.02-0.47), arthritis (e.g., Kamba (aOR 0.25 (95%CI:0.07-0.85), hypertension (e.g., Kalenjin (aOR 0.53 95%CI: 0.32-0.88), having at least one NCD (e.g., Luhya (aOR 0.52 (95%CI:0.35 - 0.79), and multiple NCDs (Kisii= aOR 0.33 (95%CI:0.11-0.96).

Participants that completed secondary or tertiary education compared with those who had completed primary, or no education had higher odds of having hypertension (aOR 1.83 (95%CI: 1.19-2.81), diabetes mellitus (aOR (aOR 1.96 (95%CI: 1.03-3.70), at least one NCD (aOR 1.58 (95%CI:1.20-2.08), and multiple NCDs (aOR 2.03 (95%CI:1.28-3.22). However, completing secondary or tertiary education was associated with lower odds of experiencing arthritis (aOR 0.54 (95%CI: 0.34-0.86) when compared with completing primary or no education.

We found that men who perceived themselves to be in moderate to good health compared with those that thought their health status was bad had lower odds of having hypertension (aOR 0.20 (95%CI: 0.10-0.38), diabetes (aOR 0.08 (95%CI: 0.03-0.24), heart disease (aOR 0.20 (95%CI:0.07-0.58), arthritis (aOR 0.17 (95%CI: 0.07-0.39), having at least one NCD (e.g., moderate= aOR 0.22 (95%CI:0.15-0.34) and multiple NCDs (e.g., good health conditions=aOR 0.10 (95%CI:0.05-0.18). However, men who perceived themselves to be in moderate (aOR 3.83 (95%CI:1.72-8.54) and good health conditions (aOR 7.38 (95%CI:3.61-15.08) compared with those that thought their health status was bad had higher odds of experiencing anxiety.

We also found that men from the middle (aOR3.26 (95%CI: 1.16 - 9.20), richer (aOR 3.50 (95%CI: 1.21 - 10.08) and richest (aOR 7.55 (95%CI: 2.67 - 21.37) indices when compared with those from the poorest wealth index; had a higher likelihood of experiencing diabetes mellitus. Likewise, men from the richest quintile when compared with those from the poorest quintile (aOR 1.59 (95%CI:1.05-2.41) were more likely to have at least one NCD. On the other hand, men from the richest quintile (aOR 0.50 (95%CI:0.26-0.94) when compared with those from the poorest quintile were less likely to experience anxiety.

Participating men who had inadequate weekly exercises (<150 min per week) compared with those with adequate weekly exercise (≥150 min per week) had a higher likelihood of having hypertension (aOR 1.51 (95%CI: 1.07-2.14), diabetes mellitus (aOR 1.90 (95%CI: 1.08 - 3.35). Similarly, participants who lived a sedentary life (sat for > 8 hours per day) compared with those who did not (≤ 8 hours per day), had higher odds of having diabetes (aOR 2.90 (95%CI: 1.30 - 6.47).

This study found that men subscribing to the other faith in Kenya compared with the Christians had higher odds of having arthritis (aOR 2.12 (95%CI:1.17-3.84) and experiencing depression (e.g., subscribing to the Muslim (aOR 3.16 (95%CI: 1.66 - 6.04) or other faith (aOR 1.52 (95%CI: 1.01- 2.26)

This study also found that living in urban settings was associated with less odds of having arthritis (aOR 0.37 (95%CI: 0.21- 0.66) and at least one NCD (aOR 0.76 (95%CI:0.59-0.97) when compared with living in the rural settings. Finally, men who had access to radio compared with those who did not (aOR 1.79(95%CI:1.02-3.13) had higher odds of experiencing anxiety.

## Discussion

The study aimed at determining the prevalence and associated factors of non-communicable diseases among men in Kenya. The study findings revealed that the overall prevalence of NCDs among men in Kenya was 9.4%, with hypertension, depression, and anxiety being the commonest conditions. These findings align with previous studies in other countries Lesotho and Napel, which showed hypertension to be the leading NCDs among men [12, 17]. Although there is a dearth of published data of the overall prevalence in other East African countries, the overall prevalence is lower than that in India of 12.4% and 15.9% of Kenyan women with at least one NCD [13, 18]. This variation in prevalence of NCD across countries could be due to differences in socioeconomic factors, healthcare access, lifestyle habits, and the age distribution of the populations [19].

In this study, hypertension emerged as the most prevalent NCD, with a significant proportion of the population affected being 3.5% which is lower than the 41.4% prevalence of the 2020 Global epidemiology study [20]. However, the low prevalence of cancer (0.1%) in this study contrasts with the increasing global burden of cancer, especially in low and middle income countries [21]. This discrepancy may reflect underreporting and the limited scope of cancer screenings in the study population.

In terms of multiple chronic conditions, the majority of the participants had diabetes and hypertension. The co-existence of diabetes and hypertension as a multimorbidity is supported by literature, as both conditions share several risk factors related to their development such as obesity, poor diet, physical inactivity, and genetic predisposition [22]. The co-occurrence of diabetes and hypertension significantly increases the risk of cardiovascular complications, including stroke, heart failure, and kidney disease [23]. Therefore, stakeholders such as ministry of health of Kenya need to sensitize masses on this multimorbidity, do early screening and treatment, and encourage lifestyle modifications interventions.

We also found a prevalent co-existence of mental and somatic NCDs. The most prevalent combination was hypertension and depression, followed closely by hypertension and anxiety. This is consistent with different studies which have reported that depression and anxiety can negatively impact blood pressure regulation resulting into hypertension [24, 25]. The frequent co-occurrence of mental health conditions with hypertension highlights the interplay between physical and psychological health [24]. Additionally, a combination of arthritis and depression was also notable among participants. The findings are consistent with other studies which reported an association between arthritis and depression [26]. Chronic pain and mobility limitations associated with arthritis often lead to reduced quality of life, social isolation, and emotional distress, making depression a common comorbidity [27]. Therefore, the government of Kenya and SSA countries need to prioritize and integrate mental health screening and support into chronic disease management.

This study also identified several factors associated with the prevalence of NCDs, including increasing age, region, residence, ethnicity, education level, perceived health status, wealth index, religion, media access, living sedentary lifestyle and physical activity. Age was statistically significantly associated with NCDs among men where men aged 25 years and older were more likely experience at least one or multiple, NCDs, including hypertension, diabetes, depression and arthritis compared with the younger participants. This concurs with the existing literature in India and Ghana, which emphasizes that aging is a key risk factor for NCDs [28, 29]. However, the finding is inconsistent with a study in Burkina Faso that reported NCDs tends to be higher in younger populations due to lifestyle factors such as increasing urbanization, sedentary behavior, and dietary shifts towards processed foods [30]. The aging process involves major physiological changes like reduced organ function and inflammation which makes an individual prone to chronic illness [31]. A lot of emphasis should be put on health educating the older population about healthy living, physical exercises and routine medical checkups.

This study found that region was statistically significantly associated with NCDs among men. Unlike anxiety, men from Northeastern, western, Rift Valley, Nairobi, and Nyanza were more likely to experience a heart disease, depression, have at least one NCD and multiple NCDs compared with those from the coastal region of Kenya. The study finding is consistent with findings from a study in Kenya which reported regional differences influence the prevalence of NCDs [7, 32]. Urban areas like Nairobi and the Coast were found to have high NCD rates due to sedentary lifestyles, unhealthy diets, and substance use, especially in informal settlements. However, in our study, less urbanized regions such as Nyanza, North-eastern, and Rift Valley also showed high NCD risk among men, which may be due to smoking, alcohol use, job-related stress, and poor access to healthcare. Even in rural areas, the growing use of processed foods is increasing NCD risk [4]. Therefore, the government should develop and implement region-specific public health policies such as the rural tobacco tax that addressed the root causes of NCDs in high-risk regions of India [33].

Tribe was statistically significantly associated with NCDs among men, as men from the Meru, Kamba, Taita/Taveta, Luhya, and Embu tribes had higher odds of experiencing NCDs like arthritis, anxiety, heart disease, and lung disease compared with the Kikuyu tribe. However, men from some ethnic tribes had lower odds of experiencing hypertension, depression, heart disease, having at least one NCD, and multiple NCDs compared with the Kikuyu tribe. The findings from this study regarding tribal differences in NCDs concur with a study in Kenya which reported the rise in NCDs in certain tribes [7]. This was attributed to increased urbanization and declining proportion of agricultural and forestry wage jobs and unhealthy eating [7]. These ethnic differences may reflect genetic, cultural, or environmental factors that influence disease susceptibility [34]. Therefore, health workers should address ethnic disparities in NCDs using culturally sensitive approaches, like the U.S. REACH program for minority diabetes care [35].

Participants that completed secondary or tertiary education compared with those who had completed primary, or no education had higher odds of having hypertension, diabetes mellitus at least one NCD and multiple NCDs, unlike experiencing anxiety. This finding concurs with previous research in India that showed a strong association between higher education levels and increased NCDs among men [36]. However, the findings are inconsistent with a study in China and a review which reported lower level of education was highly associated with hypertension and diabetes mellitus [37, 38]. Higher education in men may increase NCDs due to lifestyle changes like inactivity which is associated with white-collar jobs [39]. On the other hand, educated individuals tend to have better access to healthcare and coping strategies, which may lower their anxiety [40]. Individuals with higher education should be encouraged by various health sector stakeholders to regularly screen NCD risk factors and recognize or modify the lifestyle NCD risk factors associated with their jobs, such as long hours of inactivity, poor dietary habits, and high stress.

We also found that men with a higher wealth index, when compared with those from the poorest wealth index, had higher odds of having diabetes mellitus and at least one NCD, unlike anxiety. The findings are concur with a study in Bangladesh, which reported that diabetes risk was about 60% higher in adults from wealthy households [41]. Unfortunately, the study findings are contrary to a study in South Africa, which reported a high prevalence of diabetes among the poor [42]. Higher economic status can lead to sedentary lifestyles, stress, and unhealthy eating [43]. Wealthier individuals experience less anxiety due to better access to healthcare, financial security, and lower stress levels [44]. Health workers should prioritize educating men in higher wealth brackets the importance of maintaining a balanced diet and engaging in regular physical activity.

This study also found that participants from the urban setting when compared with those from the rural setting were less likely to have arthritis and at least one NCD. The findings concur with two studies, one in 47 counties of Kenya and another utilized data from 2022 KDHS which reported a high prevalence of cardiovascular diseases in rural areas [13, 32]. However, the study findings contrast with a study in Belagavi which reported that participants in urban areas had a high risk of NCDs and had high rate of smoking [45]. Rural areas have limited access to health care diagnostics, better transport systems, and education about healthy living. Lack of diagnostics and medical professionals delays early detection and treatment. Additionally, poor health literacy and unhealthy behaviors exacerbate NCD risks [46]. Arthritis is less common in urban settings due to better healthcare access, healthier lifestyles, and more opportunities for physical activity [47]. Policy makers in Kenya should focus on improving health awareness and early screening of arthritis in rural areas to ensure timely detection and management.

Men who had access to mass media compared with those who did not have access to it had higher odds of having lung disease, anxiety for those who had access to the radio and having internet access was associated with higher odds of having two NCDs. The findings are consistent with different studies which reported an association between access to media with lung disease, access to radio with anxiety and access to internet with having at least one NCD [18, 48, 49]. Access to radio and the internet may contribute to anxiety and unhealthy behaviors due to information overload, exposure to distressing news, misinformation, and lifestyle influences from digital media [18]. Therefore, the Kenyan government needs emphasize balanced media use and the use of communication strategies that leverage the positive aspects of media while minimizing potential risks.

Participating men who had inadequate weekly exercises compared with those with adequate weekly exercise had a higher likelihood of having hypertension and diabetes. These findings concur with a study that reported Inadequate physical activity in one-fifth of the 420 participants with hypertension [46]. Without interventions to promote regular exercise, the burden of hypertension and diabetes may continue to rise reducing overall quality of life [50]. Stakeholders should prioritize physical activity promotion as part of public health initiatives aimed at reducing the incidence of hypertension and diabetes.

Similarly, participants who lived a sedentary life compared with those who did not; had higher odds of having diabetes. The findings concur with a global study which reported that a sedentary lifestyle is associated with in-activity and poor eating habits that increase the risk type 2 diabetes [51]. This emphasizes that a sedentary lifestyle is a significant risk factor for the development of diabetes, which is a growing global health concern. Individuals should be encouraged to make small, gradual changes to reduce sedentary time and incorporate physical activity into their daily routines.

Religion was significantly associated with NCDs. Men subscribing to the Muslim or other faith compared with the Christians had higher odds of having arthritis and depression. The findings concur with a study which reported a low rate of NCDs in Christian communities in the United States due to counselling and coaching they receive to reduce NCDs [34]. Religious practices may influence health outcomes, as they often have specific health-related behaviors and social support networks that could either mitigate or exacerbate the risk of chronic diseases [52]. Stakeholders should engage minority religious communities in support groups, health education, and collaboration with religious leaders to provide early intervention and reduce stigma.

We found that men who perceived themselves to be in moderate to good health compared with those that thought their health status was bad had lower odds of having hypertension, diabetes, heart disease, arthritis, and having at least one NCD and multiple NCDs, unlike experiencing anxiety. Similar findings were observed in a study which indicated that self-rated health is a strong predictor of physical health outcomes [53]. Improving self-perception of health could lead to better health behaviors and outcomes [54]. Therefore, the government should incorporate health perception assessments into routine health screenings to motivate proactive health management and encourage behavior change.

### Strengths and limitations

We used the most-recent data from the 2022 Kenya Demographic and Health survey, with a large sample size and standardized data collection protocols. The study is limited by the fact that it is based on retrospective information provided by the survey respondents, which may be subject to recall bias. Secondly, due to a small sample size (n=8) of those who self-reported to have cancer further analysis of data related to cancer was not undertaken. In addition, the cross-sectional design of this study limits inferring causality but rather only association.

## Conclusions

This study showed a high prevalence of non-communicable diseases (NCDs) among men in Kenya, with hypertension, depression, and anxiety being the most common conditions. Factors such as age, region, residence, ethnicity, education level, health status, wealth index, religion, media access, living sedentary lifestyle and physical activity were found to be associated with having/experiencing various NCDs, reflecting both individual and environmental influences. To reduce the burden of non-communicable diseases (NCDs) among men, several targeted strategies are recommended. Older men should receive tailored health education, regular medical checkups, and physical activity promotion. Region-specific public health policies are needed to address localized risk factors, while culturally sensitive interventions should target ethnic disparities. Educated and wealthier men should be encouraged to adopt healthier lifestyles and undergo routine screenings to mitigate risks linked to sedentary jobs and affluence. In rural areas, improving healthcare access and awareness is essential. Media use should be balanced with accurate health messaging to reduce misinformation and anxiety. Promoting physical activity and reducing sedentary behavior are key to lowering hypertension and diabetes risk. Religious communities should be engaged in health promotion and stigma reduction through faith-based support and education. Finally, incorporating self-perceived health assessments into routine care can encourage proactive health behaviors and early intervention.

## Declarations

## Data Availability

Third-party data was obtained for this study from The DHS Program (https://dhsprogram.com/). Data may be requested from the DHS Program after creating an account and submitting a concept note. More access information can be found on the DHS Program website (https://dhsprogram.com/data/Access-Instructions.cfm). The data set is openly available upon permission from the MEASURE DHS website (https://www.dhsprogram.com/data/available-datasets.cfm). The authors confirm that interested researchers would be able to access these data in the same manner as the authors. The authors also confirm that they had no special access privileges that others would not have.

## Acknowledgments

We appreciate the Demographic Health Survey program for making the data available for this study.

## Funding

The authors received no specific funding for this work.

## Authors’ contributions

**Concept and proposal development**: John Baptist Asiimwe

**Data analysis:** John Baptist Asiimwe, Lilian Nuwabaine, Joseph Kawuki, Quraish Sserwanja, Angella Namulema, Grace Nambozi

**Writing the original draft:** John Baptist Asiimwe, Lilian Nuwabaine, Joseph Kawuki, Quraish Sserwanja, Angella Namulema, Grace Nambozi

**Writing- review, and editing:** John Baptist Asiimwe, Lilian Nuwabaine, Joseph Kawuki, Quraish Sserwanja, Angella Namulema, Grace Nambozi

## Ethics approval and consent to participate

High international ethical standards are ensured during MEASURE DHS surveys and the study protocol is performed following the relevant guidelines. The 2022 KDHS survey protocol was reviewed and approved by the ICF Institutional Review Board. Written informed consent was obtained from human participants and written informed consent was also obtained from legally authorized representatives of minor participants.

## Consent for publication

This is not applicable.

## Competing interests

The authors have declared that no competing interests exist.

## Notes

### Competing Interest Statement

The authors have declared no competing interest.

### Funding Statement

The author(s) received no specific funding for this work.

